# Lateralized motor control mechanisms underlie lateralized motor deficits in early Parkinson’s Disease

**DOI:** 10.64898/2026.09.09.26362567

**Authors:** Brooke Dexheimer, Caroline Selb, Jessica Manning, Narges Yaghoubi, Annalise Porreca, Matthew J. Barrett, Peter Pidcoe, Dean Krusienski, Brian D. Berman

## Abstract

**Introduction:** Parkinson’s Disease (PD) motor features typically begin unilaterally, but it remains unclear whether dominant versus non-dominant side motor symptoms differentially affect lateralized aspects of fine motor control.

**Methods:** We recruited 16 right-handed participants with mild, asymmetric PD (8 primarily left-side affected, 8 primarily right-side affected) and 10 age-matched controls to complete a bimanual tablet task in which one hand draws a stylus to targets (leveraging trajectory control mechanisms) while the other hand simultaneously stabilizes a stylus over a stationary target (leveraging stabilizing control mechanisms). The styli are connected via a resistance band to mimic internal forces that arise during asymmetric, bimanual activities of daily living.

**Results:** Both PD groups demonstrated preserved trajectory control in their dominant hand relative to controls (*p* > .072), but significantly impaired stabilizing control in their non-dominant hand (*p* < .022), with the largest deficits observed in the left-side PD group. Trajectory control performance significantly correlated with an overall measure of motor severity (*r*(14) = .69; *p* = .004), while stabilizing control did not (*p* = .56).

**Discussion:** These findings suggest that left- and right-side motor symptoms in early PD produce distinct patterns of functional impairment (rather than simple contralateral analogs) that may be related to underlying lateralized motor control mechanisms. This work highlights a potential gap in standard clinical motor assessments, which may not adequately capture non-dominant side stabilizing deficits in early PD.

## Introduction

Parkinson’s Disease (PD) is a progressive neurological disorder that affects millions of individuals worldwide each year and has sharply increased in prevalence over the past two decades [1, 2]. Individuals with PD often present with both motor and non-motor features that progressively disrupt functional independence and increase burden of care [1]. Cardinal motor features include bradykinesia, rigidity, and resting tremor [3, 4].

The cardinal motor features of PD typically begin unilaterally [5, 6], affecting either the dominant or non-dominant side at estimated rates of 57% and 43%, respectively [5]. As the disease progresses, motor features often extend their involvement bilaterally though continue to remain asymmetric in severity [6, 7]. Notably, individuals with initial non-dominant side PD appear to be diagnosed *earlier* compared to those with initial dominant side PD [5]. This difference in the interval between symptom onset and diagnosis suggests that non-dominant side PD may disrupt function more than dominant side PD in the early stages of the disease. However, it remains unclear what specific aspects of non-dominant vs. dominant side PD contribute to this difference in diagnostic latency.

Dominant and non-dominant side motor control asymmetries may contribute to differences in functional impairments during the early stages of PD. Evidence suggests that the dominant and non-dominant arms are specialized for different features of motor control [8–16]. Under this hypothesis, the dominant arm (the right arm in 90% of the population [10]) and its control system are specialized for predictive aspects of motor control that exploit intersegmental dynamics more effectively during movement planning [8, 12, 15]. This results in smoother trajectories and more energetically efficient upper extremity movements. In contrast, the non-dominant arm and its control system are specialized for impedance aspects of control that are less reliant on predictive mechanisms [9, 14]. Research in unilateral stroke has extended these findings to lateralized specializations in the dominant and non-dominant hemispheres [13, 17–21]. Individuals with unilateral dominant hemisphere lesions demonstrate contralesional and ipsilesional motor deficits in trajectory planning, while individuals with non-dominant hemisphere lesions demonstrate contralesional and ipsilesional motor deficits in impedance control [19–23].

The empirical findings reported in healthy controls and individuals with unilateral stroke suggest that unilateral motor features observed in early PD may differentially affect functional bimanual fine motor control. Specifically, non-dominant side PD may be most disruptive during aspects of control involving robust stabilization, while dominant side PD may be most disruptive during aspects of tasks involving effective trajectory control. However, the most widely used clinical measure to assess motor symptom severity, the Movement Disorder Society revision of the Unified Parkinson’s Disease Rating Scale (MDS-UPDRS), includes rapid, successive, non-functional movements that primarily involve trajectory control mechanisms, such as finger tapping and wrist pronation/supination. It remains unclear whether deficits in stabilizing control would be detected with this observational measure.

In this study, individuals with mild PD and motor feature asymmetry performed a bimanual tablet task that mimics the functional, asymmetric relationship between the hands during activities of daily living (ADLs). We predicted that individuals with primarily dominant side PD would demonstrate the largest deficits in trajectory control, especially in their dominant hand, while individuals with primarily non-dominant side PD would demonstrate the largest deficits in stabilizing, especially in their non-dominant hand. Additionally, we predicted that those with higher MDS-UPDRS Part III scores (indicating greater motor severity) would demonstrate larger deficits in trajectory control, while no relationship would be observed between Part III scores and stabilizing control, consistent with the idea that Part III scores primarily assess motor feature severity during movements involving trajectory control mechanisms.

## Methods

### Participants

We recruited 16 right-handed participants with mild PD, eight primarily right-side affected (RPD group) and eight primarily left-side affected (LPD group), from the Virginia Commonwealth University (VCU) Parkinson’s and Movement Disorders Center, as well as 10 right-handed older adult controls via convenience sampling. All participants, regardless of group, were between the ages of 55 and 90 and right-handed as determined by a score of 50 or higher on the Edinburgh Handedness Inventory – Short Form [24, 25]. Participants in the PD groups had a diagnosis of PD made by a neurologist with fellowship training in movement disorders. To isolate early-stage asymmetric motor features in PD, only participants with an MDS-UPDRS Part III score of < 35 were included [26]. Additionally, we excluded individuals who received their PD diagnosis prior to the age of 45 and/or anyone with a history of any other neurological or medical conditions that might interfere with task performance (e.g. arthritis, previous stroke, joint replacement, etc.). Control participants were individuals with no history of neurological or orthopedic conditions that might interfere with task performance. All study procedures were approved by the VCU Institutional Review Board (HM#20027379), and all participants provided written informed consent.

### Experimental Procedures

Experimental procedures are shown in **Figure 1**. All potential participants completed the following clinical assessments: 1) the Montreal Cognitive Assessment (MoCA) [27, 28], a brief cognitive assessment used to quickly screen for mild cognitive impairment, and 2) the Trail Making Test A & B (TMT-A, TMT-B) [29, 30], a fine motor and neuropsychological measure of cognitive impairment, visual scanning, mental flexibility, and executive functioning. To isolate the impact of motor deficits alone, we excluded potential participants with suspected mild cognitive impairment (MCI), defined as a MoCA score ≤ 22 (the cutoff proposed as optimal for identifying MCI in individuals with PD [31]). Additionally, motor symptoms in the PD groups were assessed using Part III of the MDS-UPDRS [32] by two separate certified raters, and administration of this assessment was video recorded. If scoring disagreements on an item arose, both raters reviewed the video recording and discussed the score discrepancy until an agreement was reached. If PD participants were being treated with dopaminergic medications and reported the presence of ‘ON’ and ‘OFF’ states within their medication regimen, the experimental procedures were conducted during their ‘ON’ state.

**Figure 1.**
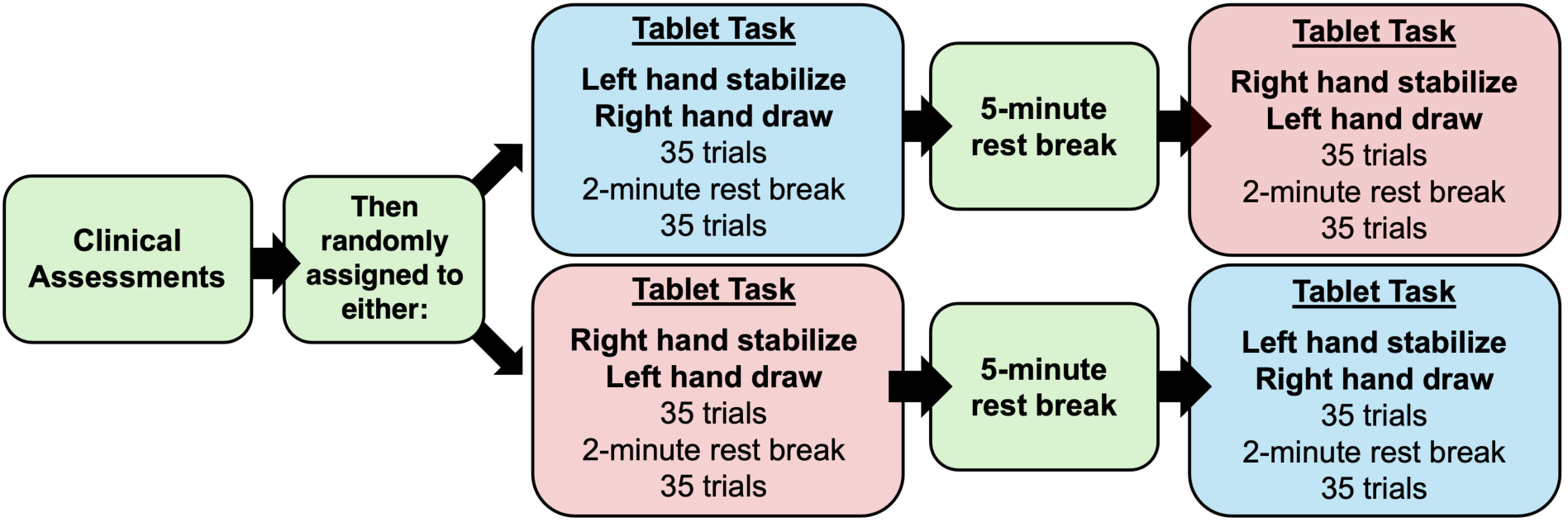
Experimental procedures

Following the clinical assessment battery, all participants completed a touchscreen tablet-based bimanual task, as shown in **Figure 2**. This task aimed to mimic the functional, asymmetric relationship between the hands during ADLs (i.e. one limb system stabilizes while the other manipulates, with both counteracting internal forces that arise). The participant was seated in a standard chair in front of a tabletop, with two touchscreen tablets (Apple iPad Pro 13”, 2020) oriented vertically and positioned side-by-side in front of them. Holding a stylus (Apple Pencil, 2^nd^ Generation, 2018) with built-up foam grips (0.25-inch radius) in each hand, participants were instructed to place each stylus tip in the respective tablet’s start circle. On one tablet, targets appeared in a pseudorandom order in one of seven on-screen locations. The participant was instructed to ‘draw’ their stylus tip from the start circle to the target as quickly and as accurately as possible. Simultaneously, in the opposite hand, they were instructed to ‘stabilize’ the stylus within the start circle on the corresponding iPad. These two styli were connected to each other with a standardized resistance band (11.2 N ± 1.4 SD at 8 cm). This resistance band was not meant to induce fatigue, but, rather, mimic the internal forces that arise during everyday bimanual activities. We considered a ‘trial’ as one ‘draw’ from the start circle to a target location, and each participant completed two blocks of 35 trials per block, with a two-minute rest break between blocks. After these 70 trials, the tablets were switched to the opposite hand configuration (see **Figure 1**), where they completed an additional two blocks of 35 trials. The hand configuration was counterbalanced between participants to minimize order effects (as shown in **Figure 1**).

**Figure 2.**
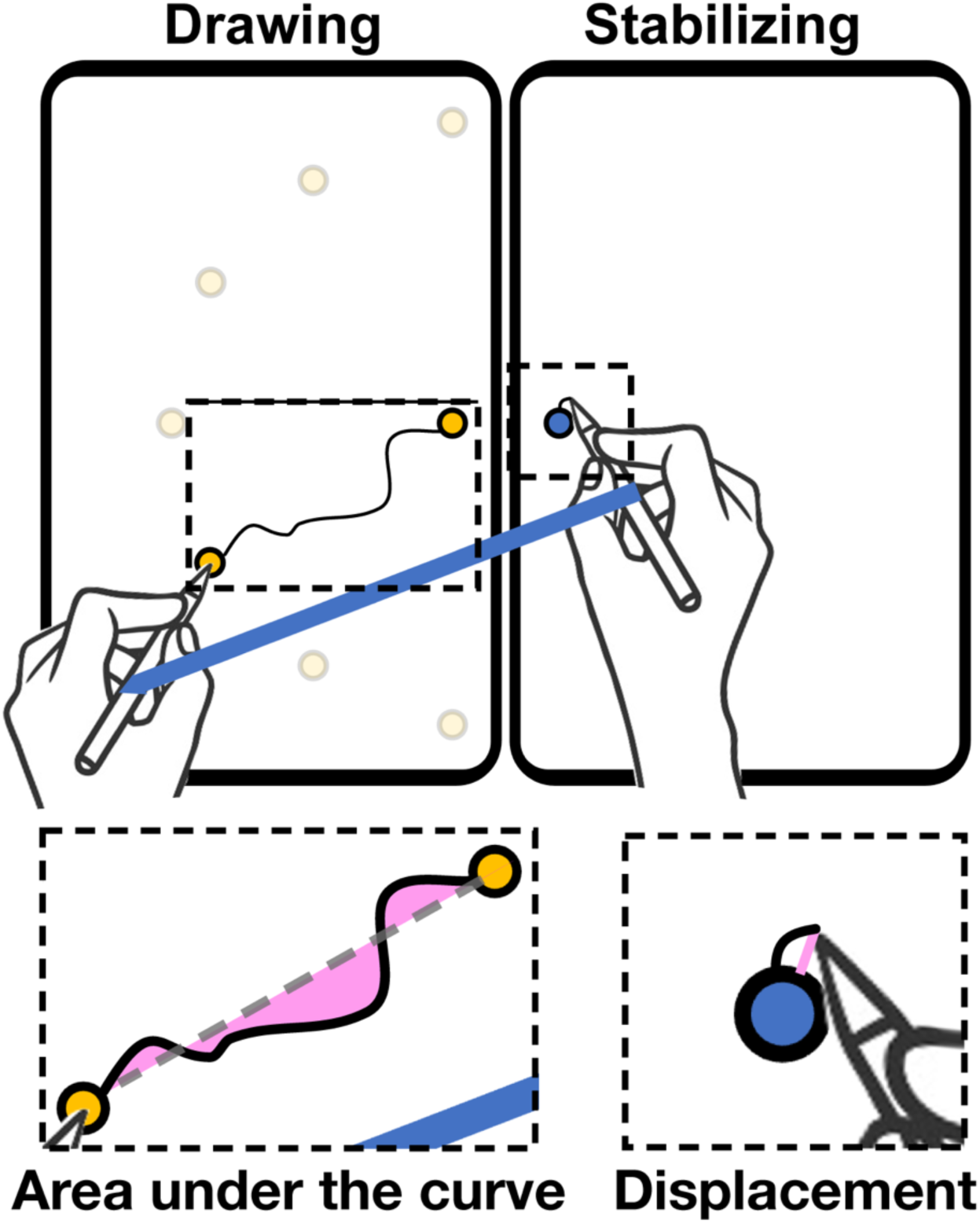
Performance measures for each portion of the touchscreen tablet-based task. During this bimanual task, participants used one hand to draw a stylus tip to targets that appeared one-at-a-time in one of seven target locations (depicted in the left tablet). Concurrently, with the opposite hand, they stabilized a stylus tip in a stationary target (depicted in the right tablet). The styli were connected by a standardized tension band (depicted in blue). Performance on the drawing task was quantified via area under the curve (pink shading, bottom left), while performance on the stabilizing task was quantified via root mean square displacement error (pink line, bottom right).

### Kinematic Data Processing

The XY coordinates from both styli were simultaneously sampled at 100 Hz using custom code written within the tablet task environment (Processing IDE; Processing Foundation [33]) and low-pass filtered using a zero-lag Butterworth filter (order: 2; cutoff frequency: 20 Hz). As shown in **Figure 2**, performance on the drawing portion of the tablet task was quantified via area under the curve (AUC), in square centimeters (indicated by the pink shading in Figure 2, bottom left). AUC was calculated by integrating the absolute perpendicular deviation between the stylus position and the ideal linear path to the target center at each sample, summed across the full time-ordered trajectory. Thus, lower AUC values indicate shorter and more linear stylus paths from the start circle to the target. Performance on the stabilizing portion of the tablet task was characterized via root mean square error (RMSE) in centimeters, calculated from the displacement of the stylus tip outside of the start circle at every 100 Hz sample collected across the full duration of the block. RMSE was selected as the primary outcome for quantifying stabilizing performance because it captures the magnitude of positional variability around the start circle, which is consistent with impedance-control demands of the stabilizing limb during bimanual ADLs. Thus, higher RMSE indicates larger positional deviations and poorer stabilizing performance.

### Statistical Analysis

We compared baseline demographics across each group using the Kruskal-Wallis test [34] (LPD vs. RPD vs. controls) and Mann-Whitney U test (LPD vs. RPD) to identify potential group differences in continuous demographic variables. Prior to analysis of performance on each portion of the bimanual task, trial-level AUC and RMSE outliers greater than four standard deviations from the hand-specific mean were removed. We then calculated hand-specific means for each participant and fit a linear mixed-effects (LME) model with hand (dominant, non-dominant) as the within-subject factor, group (LPD, RPD, controls) as the between-subject factor, and age and gender as covariates. To test group differences (LPD vs. controls, RPD vs. controls) separately for each hand, the dominant hand and non-dominant hand were each set as the reference level of the hand factor in separate parameterizations of the same LME model, yielding four pairwise contrasts. Because these are algebraic re-expressions of an identical model [confirmed by identical Akaike Information Criterion (AIC), Bayesian Information Criterion (BIC)], and log-likelihood across parameterizations), no correction for multiple model comparisons was needed; however, a Bonferroni correction was applied for the four pairwise group contrast comparisons (corrected α = .05, using an uncorrected threshold of p = .0125). Corrected p-values are reported in the manuscript body where indicated.

Additionally, we predicted that the most widely used clinical measure to assess motor symptom severity, Part III of the MDS-UPDRS, would primarily captures deficits related to trajectory control but not stabilizing control. To test this prediction, we calculated Pearson correlation coefficients to determine the predictive relationship between each individual’s overall drawing and stabilizing performance on the tablet task (summed dominant and non-dominant AUC and RMSE, respectively) and their MDS-UPDRS Part III score. Residuals were assessed for violations of normality using the Shapiro-Wilk test[35].

For all statistical analyses conducted, statistical significance was defined as an α-level ≤ .05. All analyses were performed using custom scripts written in Python version: 3.12.13 and MATLAB version: 9.13 (R2022b) [36].

## Results

### Participant demographics

Participant demographics are shown in **Table 1**. The groups demonstrated similar mean ages (*p* = .84) and MoCA scores (*p* = .48). Among the LPD and RPD groups, participants had similar MDS-UPDRS Part III scores (*p* = .24), as well as similar scores for their more-affected side (*p* = .15) and less-affected side (*p* = .39).

**Table 1.**
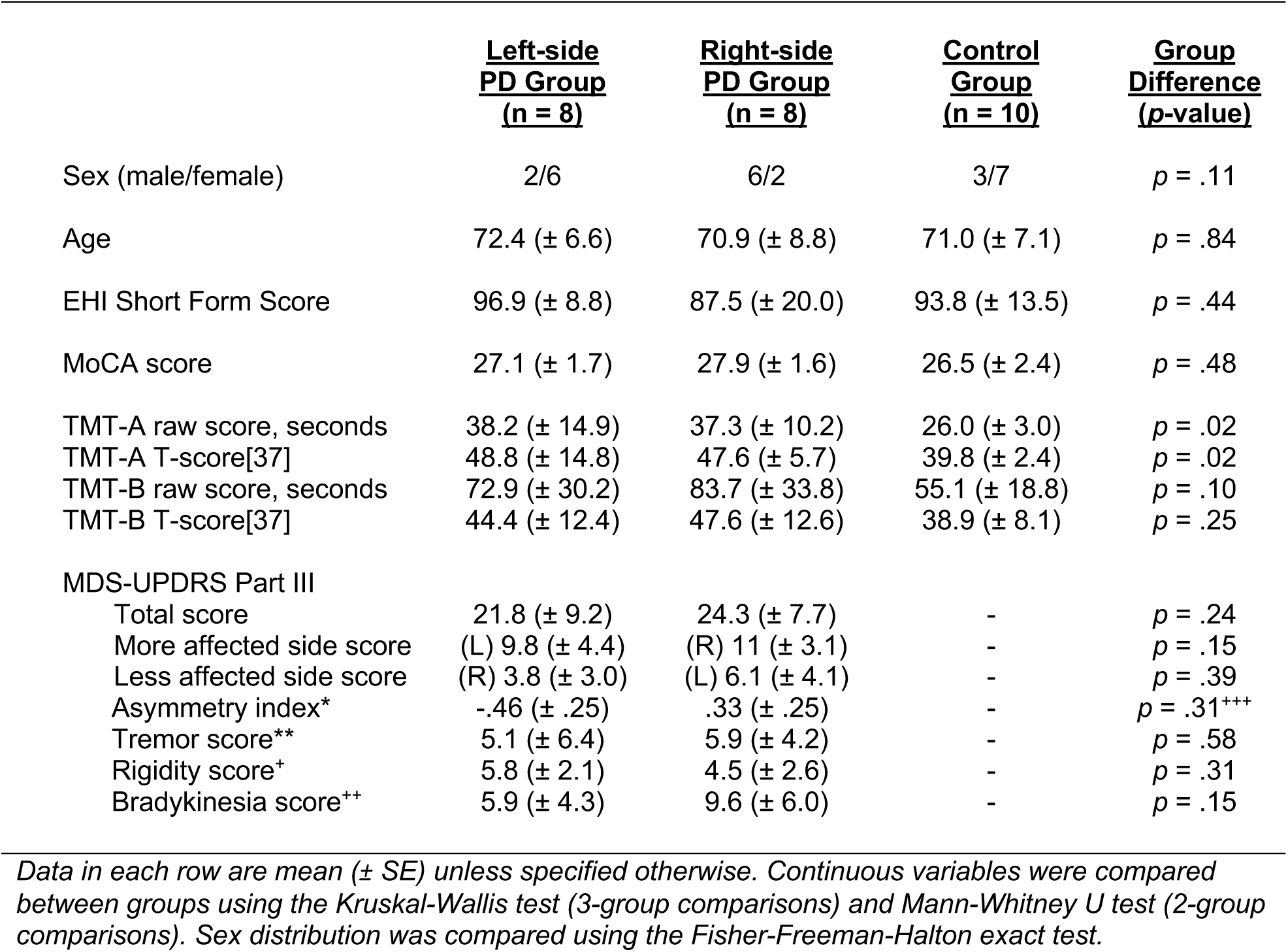
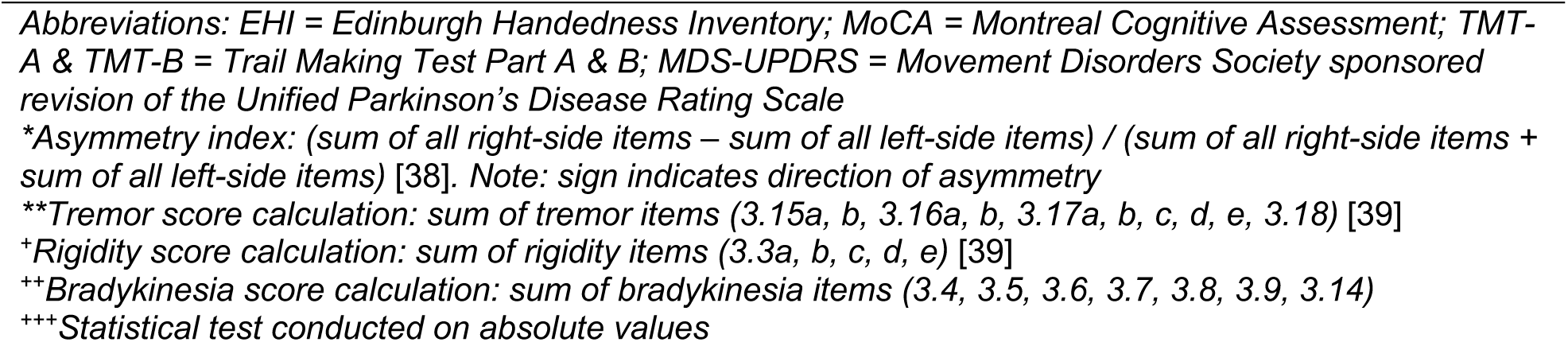
Participant demographics.

### Tablet-task performance

Stylus paths from a representative participant in each group are shown in **Figure 3**. Performance in the drawing task across the groups and hands, as measured by AUC, is shown in **Figure 4** *(left)*. Overall, participants performed with lower AUC in their dominant vs. non-dominant hand (β = 92.83, SE = 44.80, *t*(43) = 2.07, *p* = .044). Both PD groups performed similarly to controls with their dominant hand [LPD: corrected *p* = .072; RPD: corrected *p* = .513]. Conversely, in their non-dominant hand, both the LPD group (β = 231.14, SE = 81.53, *t*(43) = 2.84, corrected *p* = .028) and RPD group (β = 235.55, SE = 87.43, *t*(43) = 2.69, corrected *p* = .040) had significantly higher mean AUC than controls. Age was significantly associated with performance, with older participants showing higher mean AUC (β = 15.66, SE = 4.54, *t*(43) = 3.44, *p* = .001). Gender was also a significant predictor, with male participants performing with higher mean AUC than female participants (β = −156.62, SE = 71.87, *t*(43) = −2.18, *p* = .035).

**Figure 3.**
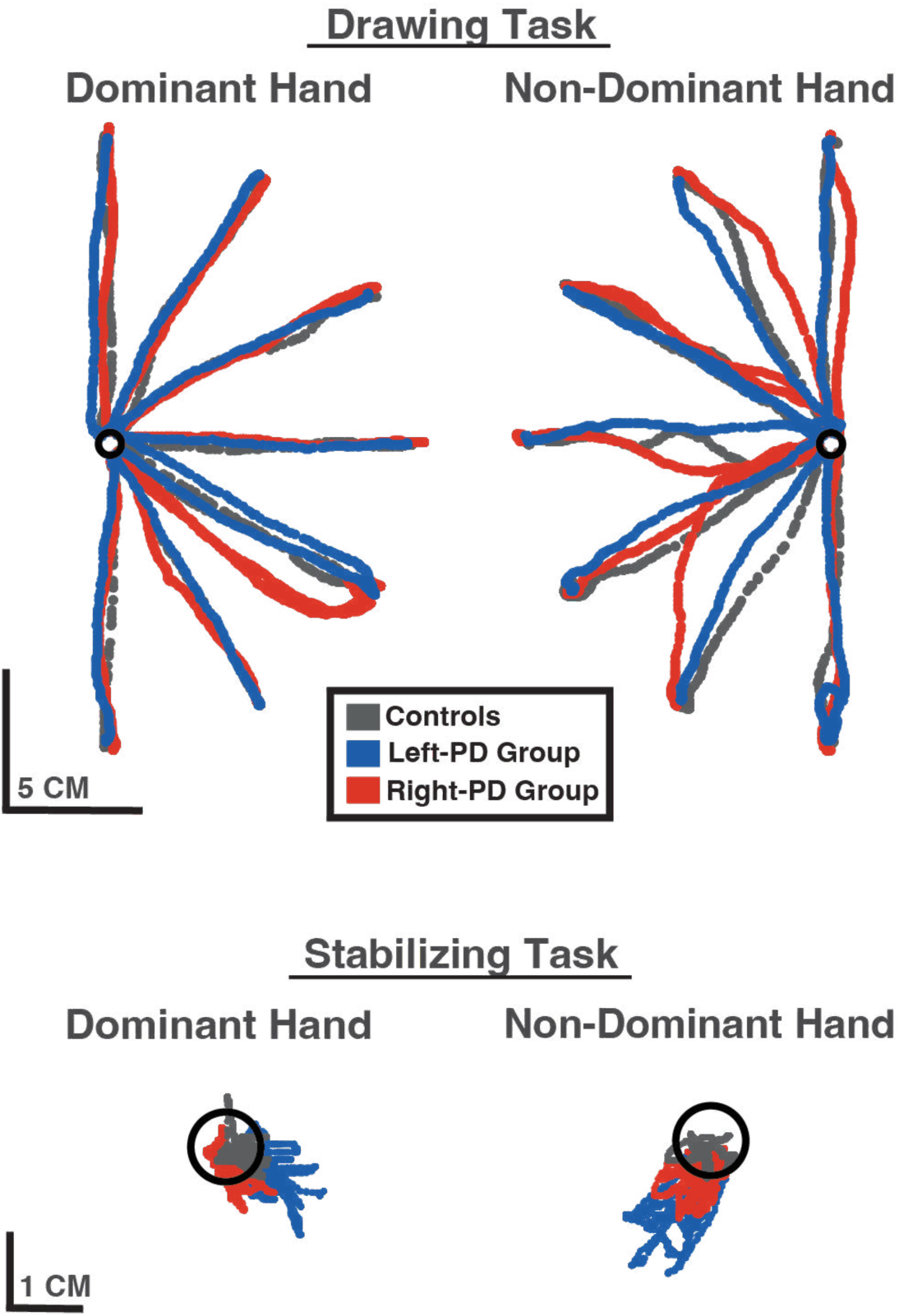
Stylus paths across both hands for a representative participant from each of the three experimental groups: older adult controls (shown in gray), individuals with primarily left-side PD (shown in blue), and individuals with primarily right-side PD (shown in red), for both the drawing (*top*) and stabilizing (*bottom*) tasks.

**Figure 4.**
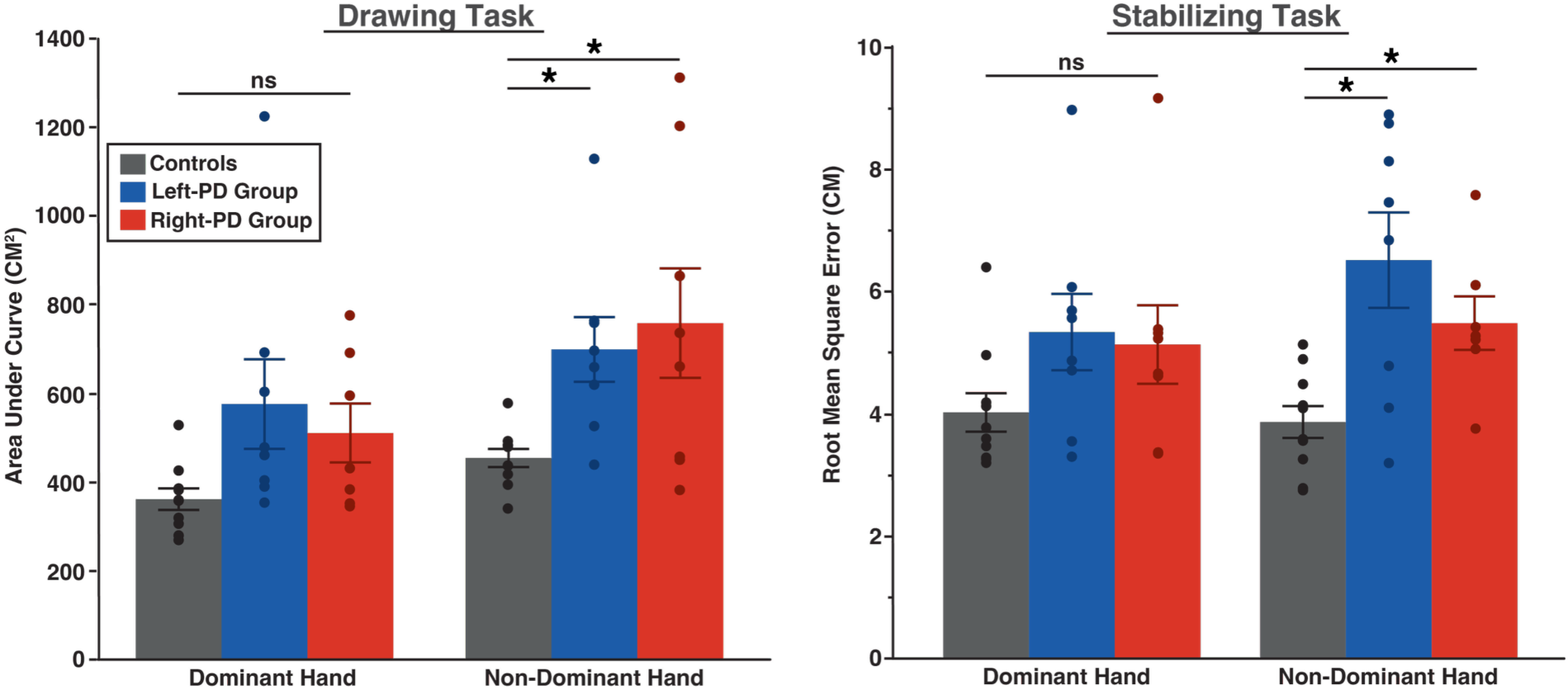
Performance differences between the LPD, RPD, and control groups for the drawing task (*left*) and stabilizing task (*right*). Both the LPD and RPD groups demonstrated preserved trajectory control in their dominant hand. Additionally, both PD groups demonstrated impaired stabilizing control, especially in their non-dominant hand, with these deficits largest in the LPD group.

Performance in the stabilizing task across the groups and hands, as measured by root mean square error (RMSE), is shown in **Figure 4** *(right)*. Overall, performance was similar between the dominant and non-dominant hands (*p* = .69). Both PD groups performed similarly to controls with their dominant hand [LPD: corrected *p* = .284; RPD: corrected *p* = .386]. In their non-dominant hand, both the LPD group (β = 2.54, SE = 0.65, *t*(43) = 3.90, corrected *p* = .0013) and RPD group (β = 2.10, SE = 0.72, *t*(43) = 2.92, corrected *p* = .022) demonstrated significantly higher RMSE than controls. Additionally, the LME revealed a significant interaction term for the LPD group (β = 1.33, SE = 0.59, *t*(43) = 2.25, *p* = .030), indicating that group differences in stabilizing performance depended on the hand used. Age was also significantly associated with performance (β = 0.075, SE = 0.036, *t*(43) = 2.10, *p* = .041), with older participants showing higher RMSE. Gender was not a significant predictor (*p* = .81).

#### Stabilizing control deficits do not correlate with clinically assessed motor severity

Performance on each portion of the tablet task was correlated with MDS-UPDRS Part III scores using Pearson correlation coefficients. As shown in **Figure 5** (*top*), MDS-UPDRS Part III scores demonstrated a significant correlation with drawing task performance (*r*(14) = .69; *p* = .004) but not stabilizing task performance (*p* = .56) (**Figure 5**, *bottom*).

**Figure 5.**
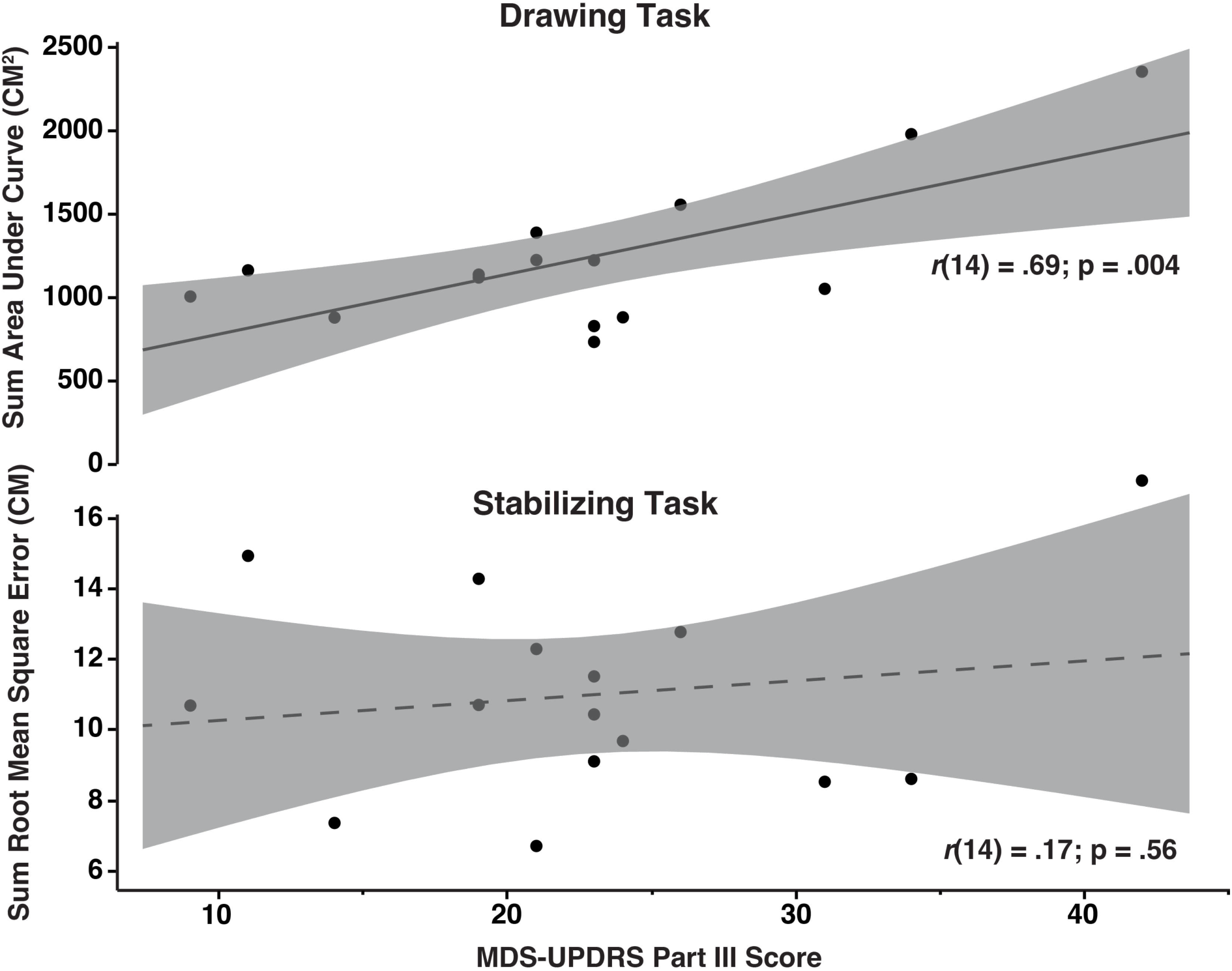
Correlations between individual MDS-UPDRS Part III score and task performance for the drawing task (*top*) and stabilizing task (*bottom*). *Note: dominant and non-dominant hand performance for each aspect of the task was summed for an overall measure of performance. One participant was excluded due to missing data. Dashed lines represent an insignificant line of fit.* Only drawing task performance was significantly correlated with MDS-UPDRS Part III score. No significant correlation was observed between MDS-UPDRS Part III score and stabilizing task performance.

## Discussion

In this study, we investigated whether early-stage left- and right-side PD motor features differentially affect lateralized aspects of functional fine motor control. While performing a bimanual task that mimics the asymmetric use of the dominant and non-dominant hands during ADLs, both the LPD and RPD groups demonstrated preserved trajectory control in their dominant hand but impaired stabilizing control in their non-dominant hand, with these deficits largest in the LPD group. This finding is in partial agreement with our overarching hypothesis that cardinal motor features of PD differentially affect functional fine motor control based on underlying specializations of the dominant and non-dominant arm control systems. Additionally, we found that trajectory control significantly correlated with overall motor severity as measured by the MDS-UPDRS Part III score, while no such correlation was observed between stabilizing control. These preliminary findings suggest that, in early stages of PD, left- and right-side motor features may not merely be contralateral analogs, but, rather, may have distinct functional impairments that depend on underlying lateralized mechanisms of motor control.

### Lateralized mechanisms of motor control may be differentially affected by the more affected side in Parkinson’s Disease

Extensive empirical findings spanning young healthy controls and numerous patient populations have suggested that the dominant and non-dominant control systems are differentially specialized for complementary aspects of motor control (see [10, 11] for review). The dominant arm and its respective control system may be specialized for energetically efficient movement that more effectively leverages intersegmental dynamics [15, 40], resulting in smoother reaching trajectories. The non-dominant arm and its respective control system may be specialized for robust stabilization due to more effective impedance control mechanisms [9, 12, 14]. This work has been extended to suggest that these lateralized mechanisms are housed in the dominant (left) and non-dominant (right) hemispheres, respectively, as right-handed individuals with unilateral stroke exhibit *bimanual* motor deficits in accordance with these proposed lateralized specializations [10, 11, 13, 18, 19, 22]. Notably, we found that those with early RPD (coinciding with potentially greater left-vs. right-hemisphere dysfunction) had preserved trajectory control in their dominant right hand compared with older adult controls, contrary to our hypothesis that these individuals would demonstrate bilateral trajectory control deficits. However, in agreement with our hypothesis, we found that those with early LPD (coinciding with potentially greater right-vs. left-hemisphere dysfunction) had impaired bilateral stabilizing control, especially in their non-dominant hand.

It remains unclear why we observed this discrepancy in deficit patterns between the LPD and RPD groups. Notably, it has been reported that those with initial motor features on their non-dominant side are diagnosed sooner relative to symptom onset [5], suggesting a unique functional disruption specific to non-dominant side PD. Thus, it is plausible that the specific contributions of the non-dominant hand to ADLs (stabilization, impedance control) may be more functionally disrupted by motor impairment in the early stages of PD. This could partially explain why the RPD group in the present study demonstrated preserved trajectory control in their dominant hand, despite being severity matched to the LPD group (which showed significantly disrupted stabilizing control in their non-dominant hand).

Alternatively, right-handed individuals with PD who are more impacted on their right side may experience a longer duration of preserved trajectory control in their dominant hand despite disease progression. This may be due to a dopamine “reserve” effect, as dopamine levels have shown to be pre-morbidly higher in the dominant vs. non-dominant hemisphere in both rats [41] and humans [42]. Because motor symptoms do not develop until ∼50% of dopaminergic neurons in the substantia nigra have degenerated [43], a higher pre-morbid baseline in the dominant hemisphere would require a proportionally greater degree of degeneration before PD motor features become functionally disruptive, which could maintain intact dominant hand trajectory control mechanisms for a longer disease duration. It has also been shown that the dominant M1 has more abundant connections, which some have suggested provides an additional compensatory mechanism for motor control following dopamine depletion [44]. Ham et al. found that, for a given concentration of dopamine transporters in the posterior putamen, those with greater non-dominant side symptoms had worse overall motor severity scores than those with greater dominant side symptoms [44]. Thus, in the present study, these asymmetries could partially explain why the RPD group had preserved trajectory control despite being severity-matched to the LPD group. However, because we did not restrict participation to solely drug-naïve PD participants, additional investigation is necessary to determine how dopaminergic asymmetries between the hemispheres might contribute to the findings in the present study and the dominant vs. non-dominant side PD diagnosis timeline discrepancy reported by Barrett et al. [5]

### Do alterations in shifting motor attention contribute to bimanual motor control deficits in early PD?

Previous literature has demonstrated bimanual fine motor deficits in PD during tasks requiring different roles of the left and right hands [45–47], similar to the bimanual tablet task used in the present study. These deficits may not solely be due to PD motor features but, rather, could be partially due to downstream disruptions in motor control and motor attention mechanisms. In support of this hypothesis, Horstink et al. had participants with PD and older adult controls perform two separate tasks simultaneously: drawing with the dominant hand while rhythmically squeezing a bulb with the non-dominant hand [46]. They found that PD patients had preserved drawing function and squeeze frequency when compared to controls, but diminished squeeze amplitude. These findings suggested that while ‘time-sharing’ between the dominant and non-dominant control systems was intact, the mechanisms contributing to attention shifts were disrupted (because squeeze amplitude, which they considered a spatial aspect of the task, was disrupted). Further supporting this hypothesis is the finding that PD patients perform asymmetric simultaneous movements in a more sequential fashion (i.e. performing with one hand, pausing, then performing with the opposite hand) [47]. Thus, it is possible that the non-dominant hand drawing and stabilizing deficits observed in our PD groups could partially be due to alterations in shifting motor attention in early PD, although further research is necessary to directly test this hypothesis.

It is worth noting that the non-dominant hand trajectory and stabilizing control deficits observed in the RPD and LPD groups could also be due to disruptions in motor control mechanisms beyond just motor attention. Loehrer et al. had PD patients complete a bimanual fine motor task (finger tapping in one hand with concurrent finger button pressing combinations in the other) [45]. Using diffusion-weighted imaging, they found that white matter abnormalities - some left-lateralized (anterior thalamic radiation, temporal-occipital fasciculi, hippocampal regions) and some bilateral (superior longitudinal fasciculus, corticospinal tract, cerebellum, cingulum) - predicted the degree of bimanual impairment. However, because they did not distinguish between predominately left- and right-sided PD [45], it remains unclear how lateralized mechanisms of bimanual control were distinctly affected. Taken together with our current study, these findings support the idea that individuals with PD have bimanual fine motor deficits in both trajectory and stabilizing control that may not be solely due to the presence of PD motor features. It is plausible that additional partial contributors to these deficits during asymmetric fine motor control are disruptions in lateralized mechanisms of both motor control and cognitive processes, such as the mechanisms contributing to shifting attention between the limbs.

### Non-dominant side assessment and intervention in early Parkinson’s Disease

In the present study, we found that drawing performance on the bimanual tablet task significantly correlated with overall motor severity as measured by the Part III score, while stabilizing performance did not, despite both groups showing significant stabilizing deficits. These findings were consistent with our prediction that those with greater motor severity would demonstrate larger deficits in trajectory control, while no relationship would be observed between Part III scores and stabilizing control, consistent with the idea that Part III primarily assesses motor feature severity during movements involving trajectory control mechanisms. Because Part III is a global measure of motor severity that also captures lower extremity and axial motor deficits, this correlation should be interpreted as a relationship with overall motor severity rather than a specific upper extremity motor deficit. Our sample size did not support meaningful subscale-level analysis, and included participants had early PD that might not yet show enough variability across the 0-4 range scored on each Part III item. Nevertheless, a question that arises is whether lateralization of PD motor deficits is resolutely captured in current motor assessments, especially with regard to functional impacts. Future research is necessary to determine how movements that largely rely on stabilizing control mechanisms affect PD motor feature presentation, especially in non-dominant side PD. Importantly, some have suggested that subtle motor deficits may be an earlier clinical sign of PD, such as slight changes in handwriting velocity and height [48]. Whether or not stabilizing deficits could be an early clinical sign in non-dominant side PD requires longitudinal research in the prodromal or newly diagnosed stage onward, as well as larger, cross-sectional samples stratified by hand dominance and side of symptom onset.

Finally, the motor asymmetries in early-stage PD identified with our task might have relevance for more tailored neurorehabilitative interventions in occupational or physical therapy. In rats, greater frequency of limb use appears neuroprotective against nigral damage[49], and, based on these findings, some have suggested that more frequent use of the dominant upper extremity might provide a protective factor in individuals with early-stage PD [44]. However, it remains unclear whether neurorehabilitative interventions specifically targeting stabilizing mechanisms are preferable to more general motor interventions for maintaining non-dominant hand function in PD. Presently, the opposite has been suggested: rigorous physical activity has global protective factors against PD progression that are irrespective of lateralized motor control mechanisms [50, 51] (see [52] for review). Future research should determine whether bimanual asymmetric assessments and interventions, such as the bimanual tablet task used in the present study, can inform more individualized neurorehabilitation. For example, it remains unclear whether LPD and RPD patients might benefit differentially from stabilization-focused vs. general motor interventions in each hand.

### Current study limitations

There are several limitations of this study that warrant follow-up research. We recruited a small convenience sample for this study; therefore, follow-up research with larger, more heterogenous sample sizes are necessary. Additionally, future research could delineate how specific cardinal motor features of PD might differentially affect lateralized motor deficits. For example, it remains unclear specifically how more severe bradykinesia, tremor, or rigidity might selectively hinder drawing and stabilizing performance in the dominant and non-dominant hands. In our PD cohort, participants demonstrated mild motor feature severity, so we were unable to delineate specific relationships between severity of each distinct motor feature and an individual’s drawing and stabilizing performance. Additionally, we did not collect each participant’s levodopa equivalent daily dose (LEDD), relying instead on MDS-UPDRS Part III scores to characterize only motor severity. Thus, we cannot rule out the possibility that variability in medication burden contributed to our findings. Participants in our cohort were also tested in their ‘on’ state, if applicable. As such, motor symptom fluctuations could have affected performance on either aspect of the tablet task, as dopaminergic treatment in PD does appear to improve unimanual coordination [53]. However, in a recent study using a bimanual task mimicking jar opening, medication was not found to affect average load force and temporal features of the task [54], although PD patients still exhibited abnormal force production relative to controls. Future research should more tightly constrain medication timing and LEDD and/or assess performance only in drug-naïve individuals to delineate exact medication contributions to both trajectory and stabilizing control.

## Conclusion

Motor symptoms in PD often begin unilaterally, and this study provides preliminary evidence that early-stage primarily left- and right-side motor symptoms in PD produce distinct patterns of functional bimanual fine motor impairment rather than simple contralateral mirror images. Both PD groups showed preserved dominant hand trajectory control but impaired non-dominant hand stabilizing control, with the largest deficits in the left-side PD group. This pattern is consistent with a model in which the dominant and non-dominant control systems rely on distinct, lateralized mechanisms of motor control. Furthermore, stabilizing deficits did not correlate with MDS-UPDRS Part III scores, suggesting a possible gap in identifying functionally relevant motor asymmetries in early PD. These preliminary findings support the broader hypothesis that lateralized motor control specializations, and not motor impairment alone, shape functional fine motor performance in early PD. Larger, adequately powered studies are needed to confirm these patterns, clarify their distinct neural mechanisms, and evaluate whether functional and clinical assessments of motor asymmetry should be considered separately in diagnosis and treatment planning for individuals with early PD.

## Data Availability

All data produced in the present study are available upon reasonable request to the authors.

